# Associations between maternal infections during pregnancy and childhood IQ scores using the ALSPAC birth cohort

**DOI:** 10.1101/2022.03.09.22272147

**Authors:** Janell Kwok, Hildigunnur Anna Hall, Aja Louise Murray, Michael Vincent Lombardo, Bonnie Auyeung

## Abstract

Maternal prenatal infections have been linked to children’s neurodevelopment and cognitive outcomes. It remains unclear, however, whether infections occurring during specific vulnerable gestational periods can affect children’s cognitive outcomes. The study aimed to examine maternal infections in each trimester of pregnancy and associations with children’s verbal, performance, and total IQ scores. The ALSPAC birth cohort was used to investigate associations between maternal infections in pregnancy and childhood IQ outcomes. Infection data from mothers and cognition data from children were included with the final study sample size comprising 7,410 mother-child participants. Regression analysis was used to examine links between maternal infections occurring at each trimester of pregnancy and children’s cognitive IQ scores at 18 months, 4 years, and 8 years. Infections in the third trimester were significantly associated with decreased verbal IQ at age 4 (*p*<.05, adjusted R^2^ = .004); decreased verbal IQ (*p*<.01, adjusted R^2^ = .001), performance IQ (*p*<.01, adjusted R^2^ = .0008), and total IQ at age 8 (*p*<.01, adjusted R^2^ = .001). Results suggest that later maternal infections could have a latent effect on cognitive development, only emerging when cognitive load increases over time, though magnitude of effect appears to be small. Performance IQ may be more vulnerable to trimester-specific exposure to maternal infection as compared to verbal IQ. Future research could include examining potential mediating mechanisms on childhood cognition, such as possible moderating effects of early childhood environmental factors, and if effects persist in future cognitive outcomes.

## Background

Research has suggested links between a mother’s immune system during pregnancy and fetal growth and brain development (Alvarado et al., 2017; Bale, 2009). Infections in pregnancy, such as herpes simplex virus and rubella, have been associated with intrauterine growth restriction, prematurity, and long-term neurodevelopmental deficits or disability (Curcio et al., 2020). Some evidence suggests that effects of infection on child development depend on pregnancy trimester. Maternal prenatal bacterial infections are linked with lower mean IQ scores at age 7 (Carter et al., 2003), and viral infections have shown associations with decreased IQ scores at age 7 with strong effect sizes (Lee et al., 2020), as well as with learning disabilities (Racicot et al., 2017). However, in general, the impact of infection during pregnancy on cognitive outcomes has been under-explored. In particular, it is not clear whether cognitive outcomes of infection depend on the trimester of infection, whether they impact cognitive development in different domains similarly, and whether the effects persist throughout child development. Therefore, a high-quality longitudinal cohort sample can help to elucidate associations between maternal infections occurring at specific trimesters and cognitive outcomes in childhood.

The occurrence of viral maternal infection is proposed to influence the pathogenesis of infant central nervous system abnormalities via placental transfer-mediated vertical transmission (Silasi et al., 2015). This transmission may further disrupt the fetus’ organ and brain tissue development, negatively affecting the infant’s immune system, resulting in further pathology of a compromised immune system and increased risk of childhood inflammation (Bale, 2009). Childhood low-grade inflammation then activates a systemic immune response associated with negative effects on brain development and function, with research showing with small to moderate effect sizes on general intelligence (Mackinnon et al., 2018). These processes reflect multiple potential mechanisms of harm acting upon the child when maternal infection is present during pregnancy.

Even though trimester-specific associations have been reported between maternal infections and fetal neurodevelopment (Lee et al., 2020), evidence is still lacking on whether cognitive outcomes depend on the specific timing of maternal infection exposure during gestation. The idea of fetal neurodevelopment being stage-dependent is based on the fact that brain myelination only begins during the second trimester of pregnancy with further growth reinforcements in the third trimester, followed by postnatal growth (Cordeiro et al., 2015; Yarnykh et al., 2018). Maternal inflammation such as infections during specific gestation periods may differentially affect the fetal growth process. However, there are currently a limited number of studies linking maternal inflammation occurrence in later gestational periods with negatively effects on a child’s cognitive-developmental infrastructure (Meyer et al., 2006) and functional connections such as working memory (Rosenberg, 2018). In the context of maternal inflammation and child IQ outcomes, differential effects on performance or verbal IQ have not been examined in literature.

Current research shows converging evidence between animal and human studies, with a review of large cohort istudies suggesting a causal relationship between prenatal maternal inflammation and child outcomes (Hantsoo et al., 2019), particularly with effects on the fetal brain development (Lautarescu et al., 2020). Examining the links between prenatal infection at different stages of gestation and cognitive outcomes is, however, subject to significant methodological challenges. In particular, it is not possible to conduct randomized experiments in humans for obvious ethical reasons. This makes it difficult to attribute causality to a factor such as prenatal infection. On the other hand, animal studies in which experimental manipulations are possible have been criticized for potential problems with generalization to humans (Harvey & Boksa, 2012). As such, the field must rely ontriangulation across these different evidence sources with complementary strengths and weaknesses (Banik et al., 2017; Morales-Suárez-Varela et al., 2010; Mulkey et al., 2019).

Despite the fact that human studies must rely on observational evidence, they can provide valuable predictive information about which children may be most at risk of poorer cognitive outcomes. For example, identifying an association between prenatal infection at a particular stage of pregnancy and poorer cognitive ability can help identify children who may benefit from a higher level of monitoring and intervention early in life, such as screening for learning difficulties or prioritization for early-life preventive interventions. This study seeks to utilize observed evidence to provide predictive evidence of mother-fetal associations, despite limitations in examining specific underlying processes or mechanisms.

The aim of this study is to use a cohort sample to investigate whether the occurrence of maternal infections in each trimester of pregnancy is associated with children’s verbal, performance, and total IQ scores.

## Design

### Setting

Data for this project were from the Avon Longitudinal Study of Parents and Children (ALSPAC), a population-based cohort study based in Avon, United Kingdom (UK), focusing on pregnant women with expected dates of delivery 1st April 1991 to 31st December 1992 (Boyd et al., 2013; Rosenberg, 2018). Regular follow-up clinic assessments were conducted over the years to track development over time. The ALSPAC dataset is a large, nationally representative sample of parents and children from the UK. Detailed medical information were gathered from pregnant women and their children from 8 weeks of gestation up to 22 years of age. Participants in ALSPAC have a slightly higher sociodemographic profile on average as compared to the general population in Avon and Great Britain (Fraser et al. 2013). For example, ALSPAC mothers are more likely to be married, live in owner-occupied accommodation, have a car in their household, and less likely to be non-White.

### Participants

The final sample of data collected and used in this study for when the children were seven years old was n=15,645. For the purposes of this study, participants with missing infection data across all trimesters (n=2,125) were excluded from the sample, as well as those who had missing outcome data across all cognitive domains (n=6,110). The final sample size comprised n=7,410 mother-child participants.

### Outcome measures

## Prenatal infections

Data on prenatal infections from the first, second and third trimesters were gathered in weeks 18 and 32 of gestation, and 8 weeks post-partum (Fraser et al., 2013). Retrospective assessment was completed at 8 weeks after birth for infections occurring in the third trimester of pregnancy. Women were asked whether they experienced any of the following infections: urinary tract infection, influenza, rubella, thrush, genital herpes, or other infections. Response options provided at 18 weeks were: ‘Yes, in 1st to 3 months’, ‘Yes, 4 months to now’, ‘Yes for both time periods’ and ‘Not at all’. Response options at week 32 and 8 weeks postpartum were: ‘Yes’, ‘No’, and ‘Don’t Know’. Response options for infections reported occurring during the first, second, or third trimester of pregnancy were coded and termed as ‘any trimester’. Infection data were coded into dichotomous variables, where any incidence of infection was coded as a ‘yes’, and no incidence was coded as a ‘no’. Infections were further coded according to trimesters, with any occurrence of infection coded as present (e.g., rubella or urinary tract infection at 18 weeks = infection present in the first trimester).

### Childhood IQ scores

Cognitive measures included the Griffiths Mental Development Scales (GMDS), administered at 18 months, the Wechsler Preschool and Primary Scale of Intelligence – Revised UK edition (WPPSI-RUK), administered at 4 years, and the Wechsler Intelligence Scale for Children, 3rd edition (WISC-III), administered at 8 years. The GMDS evaluates early development and consists of five subscales (locomotion, personal/social skills, hearing and speech, hand-eye coordination, and performance) (Griffiths, 1970). The hearing and speech scale (language, response, speech development) (Rosner & Simon, 1971) and the performance scale (skill development and manipulation), comprising 86 items each, were selected for use in the current study. Scores were converted to standardized developmental quotients by the ALSPAC team, with scores ranging from 0 to 100 points. An average GMDS development score variable was also included as part of the overall GDMS developmental assessment by the ALSPAC team. All variables were age-adjusted by the ALSPAC research team. The study further included two measures of cognitive functioning: WPPSI-RUK for children aged 3-7 years (Wechsler, 1989) and a short-form version of the WISC-III for children aged 6-17 years (Wechsler, 1991; Wechsler, 1993). Both tests comprise two scales: verbal and performance (non-verbal ability and executive function), with each scale containing five subtests. Scores on both the WPPSI-RUK and the WISC-III range from 40 to 160. All IQ scores were used as continuous variables. Descriptive statistics for verbal IQ, performance IQ, and fullscale IQ scores are provided in Table 1.

**Table 1.**
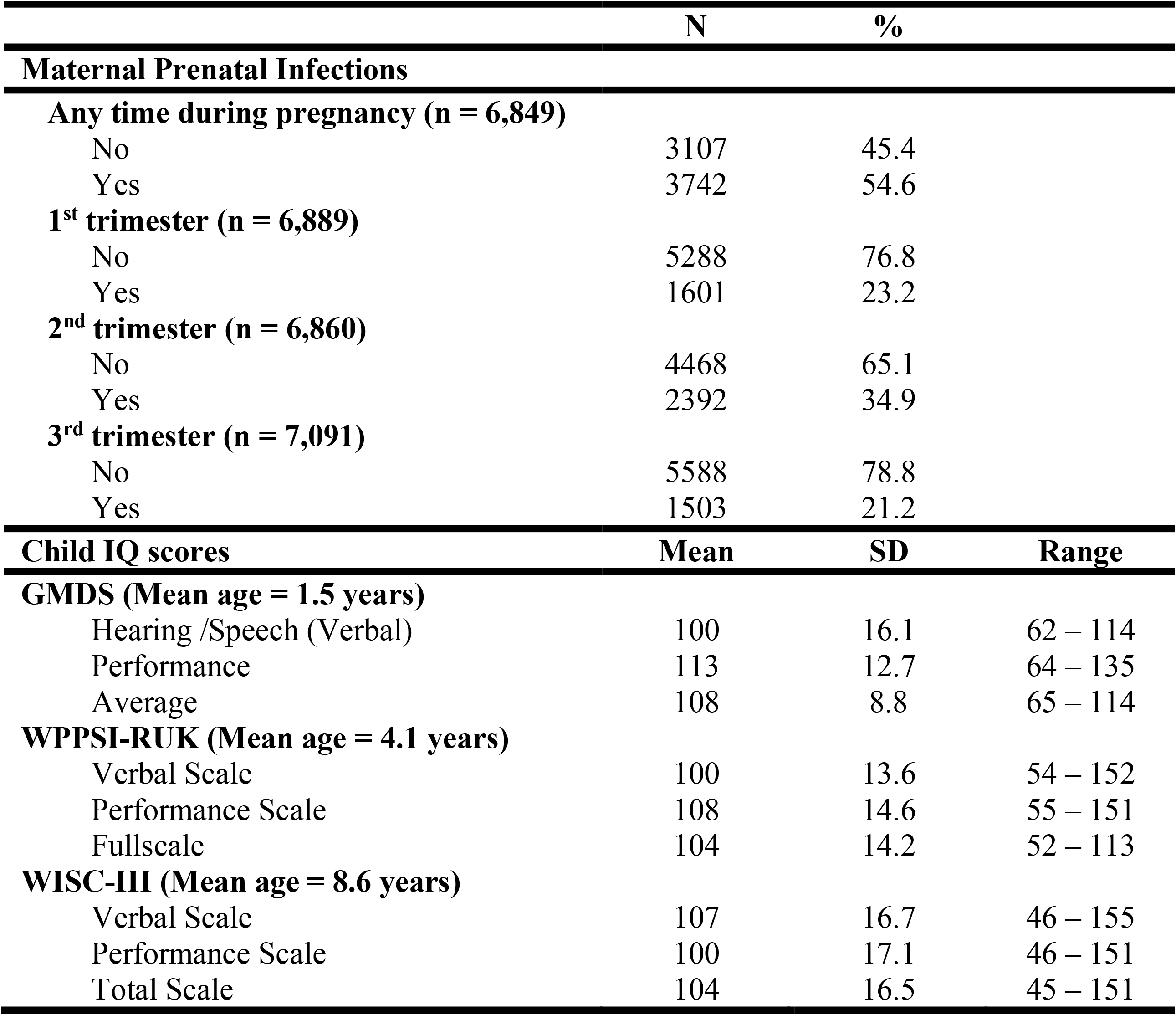
Descriptive statistics for predictors and outcomes

### Confounders and Covariates

Data on other perinatal and social factors which may be associated with prenatal infections and/or children’s cognitive outcomes were gathered from assessments from mothers and children. Child factors included child sex (male/female), gestational age (weeks) and birthweight (grams). Standard cutoffs for prematurity were applied to gestational age (≥ 37 weeks/<37 weeks) and birthweight (≥ 2500g/<2500g) as per clinical standards of falling within the 10th percentile of pregnancies, with increased infant mortality risk (Eskes et al. 2019; Hughes, et al. 2017). Maternal and socioeconomic factors included maternal age at birth, maternal education, maternal history of smoking, maternal psychiatric history, and deprivation indices. Maternal education was assessed at 32 weeks of gestation through interviews and categorized as A-levels or lower. Maternal smoking was coded as yes or no and assessed through questionnaires for every trimester of pregnancy (18 weeks, 32 weeks, 8 weeks postpartum for the third trimester). Maternal psychiatric history was assessed in the first trimester, where women were asked if they had any psychiatric conditions (anorexia, bulimia, severe depression, schizophrenia, other psychiatric issues), and coded as yes if they indicated having any psychiatric illness. Deprivation indices were sorted into quintile ranks and measured by the official Indices of Multiple Deprivation, an area-level deprivation index measured in England accounting for several domains of deprivation: income, employment, health, education, housing, and living environment (Noble et al., 2006).

Previous literature has identified these factors to be associated with children’s cognitive development (Mackes et al., 2020; Sania et al., 2019; Sommerfelt et al., 2000). These factors were classified into potential confounders and covariates for the regression models in this study, with confounders being selected based on whether previous literature showed associations between infection exposure and cognitive outcomes (such as deprivation), and covariates (such as child’s sex) selected based on their potential association with children’s cognitive outcomes. Adjusting for the latter was of interest to determine the unique contribution of infection exposure over and above other established influences.

### Statistical Analysis

All statistical analyses were conducted using R (version 3.4.1) (R Core Team, 2013). Data was inspected for and met parametric statistical assumptions. Linear regression models were used to examine the relations between infections occurring at each trimester and children’s cognitive scores at ages 18 months, 4 years, and 8 years. Three types of models were fit: an unadjusted model, a model adjusted for confounders, and a model adjusted for confounders and additional covariates. These models were run in a hierarchical manner, with increasingly adjusted models specified by adding variables to models from previous steps. The fully adjusted model included possible confounders of maternal and socioeconomic factors, and the additional covariates of child sex, gestational age, birthweight, maternal psychiatric history, and maternal smoking (Table 2). Estimates for infections in each trimester were also adjusted for infections in the other two trimesters.

**Table 2.** Confounders and covariates (n = 7,410)

| **Confounders** |  |  |
| --- | --- | --- |
|  | **Mean (SD)** | **Range** |
| **Maternal age at childbirth (years)** | 29.1 (1.23) | 15 – 44 |
|  | **N** | **%** |
| **Maternal education** |  |  |
| (≥ A' levels) | 3068 | 41.3 |
| (< A' levels) | 4111 | 55.5 |
| Missing | 231 | 3.2 |
| **Deprivation index** |  |  |
| Highest quintile | 1965 | 26.5 |
| 60-80% | 1162 | 15.7 |
| 40-60% | 1054 | 14.2 |
| 20-40% | 885 | 11.9 |
| Lowest quintile | 765 | 10.3 |
| Missing | 1579 | 21.3 |
| **Covariates** |  |  |
| **Child Gender** |  |  |
| Male | 3740 | 50.4 |
| Female | 3670 | 49.6 |
| **Child Gestation** |  |  |
| ≥ 37 weeks | 6982 | 94.2 |
| < 37 weeks | 428 | 5.8 |
| **Child Birthweight** |  |  |
| ≥ 2500g | 6970 | 94.0 |
| < 2500g | 349 | 4.7 |
| Missing | 91 | 1.3 |
| **Maternal psychiatric history** |  |  |
| No | 5955 | 80.4 |
| Yes | 829 | 11.2 |
| Missing | 626 | 8.4 |
| **Maternal smoking (any point in pregnancy)** |  |  |
| No | 5073 | 68.5 |
| Yes | 1488 | 20.1 |
| Missing | 849 | 11.4 |

In the primary analysis, missing data were handled with full information maximum likelihood estimator, and an alpha level of 0.05 indicated statistical significance (p<.05). Sensitivity analysis included multiple imputation models using the MICE package in R, with 25 imputations = (van Buuren, 2012; Royston, 2004). Models were fit from the multiple imputed data sets, with estimates pooled into a single set of estimates and standard errors, which was then used for the analysis.

## Results

Descriptive statistics showed that the occurrence of maternal prenatal infections at any time during pregnancy was 54.6%, with most infections occurring during the 2nd trimester (34.9%) (Table 1). Mean IQ scores were similar to the general population (GMDS = 108, WPPSI-RUK = 104, WISC-III = 104) (Table 1). Table 2 shows further descriptive statistics for maternal variables and children’s birth outcomes.

Unadjusted models found no links between infections occurring in the first or second trimesters and children’s outcomes. Significant associations were found between third trimester infections and WPPSI-RUK verbal scores (β= -.076, *p*=.031) at age 4, and WISC-III verbal IQ scores (β= -.036, *p*=.008), performance IQ scores (β= -.036, *p*=.009), and total IQ scores (β= -.040, *p*=.003) at age 8 (Table 3). These results showed that an infection during the third trimester of pregnancy was associated with a 2.5 IQ point decrease for WPPSI-RUK verbal scores at age 4, 1.5 IQ point decrease in WISC-III verbal scores, 1.5 IQ point decrease for WISC-III performance scores, and 1.6 IQ point decrease for WISC-III total scores at age 8. No significant associations were found between prenatal infections in all three trimesters and any of the children’s IQ outcomes (Model 1).

**Table 3.**
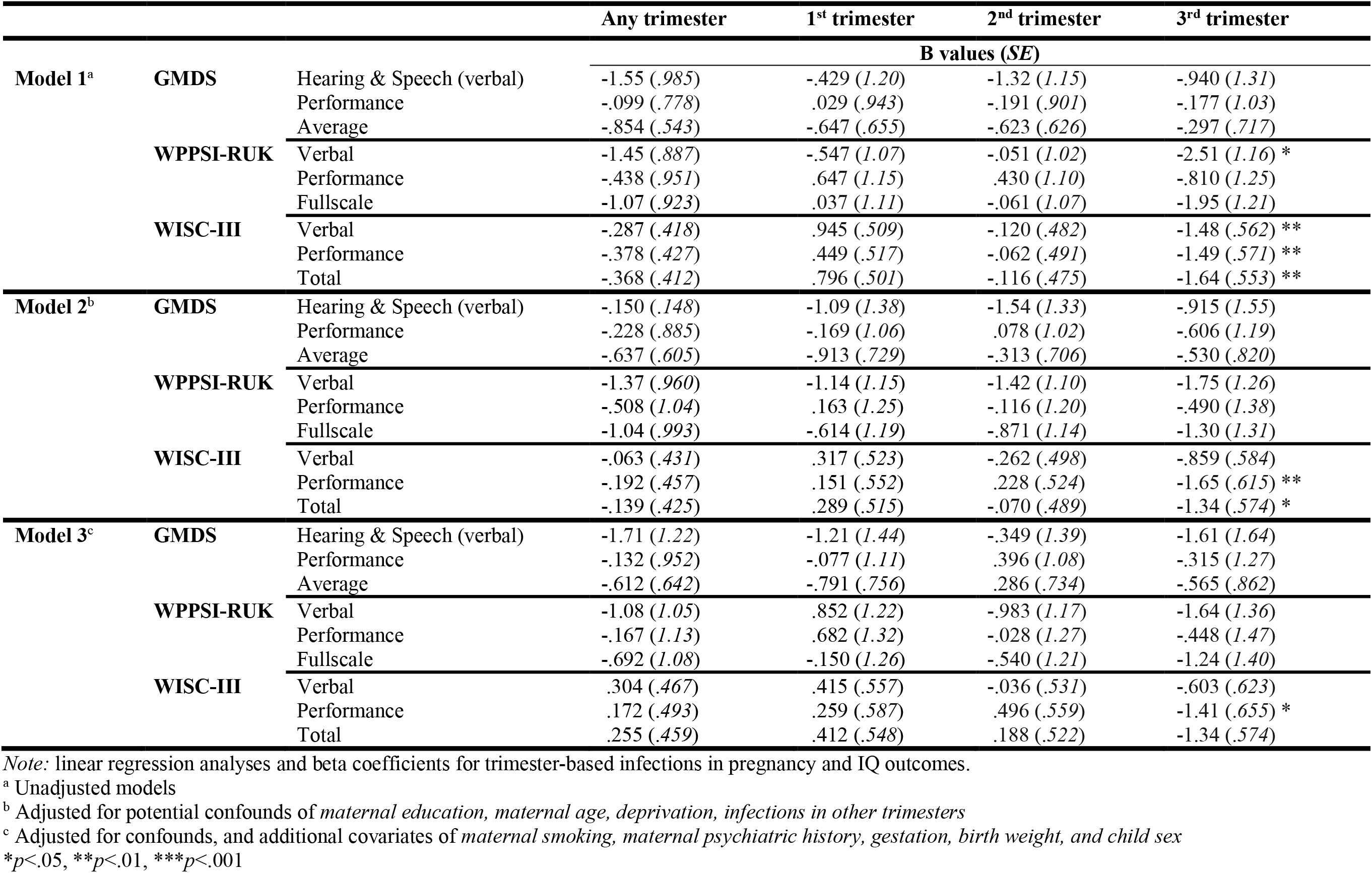
Hierarchical regression models on trimester-based infections in pregnancy and cognitive outcomes (n = 7,410)

Significant associations were attenuated after the first adjustment for confounders maternal age, maternal education, deprivation, and infections in the other two trimesters (Model 2). Associations were no longer found for WPPSI-RUK verbal IQ scores (β= -.053, *p*=.166) at 4 years and WISC-III verbal IQ scores (β= -.021, *p*=.141) at 8 years. Significant associations remained between third-trimester infections and WISC-III performance IQ scores (β= -.039, *p*=.007) and WISC-III total IQ scores (β= -.033, *p*=.020) at 8 years (Table 3). These results showed that an infection during the third trimester of pregnancy was associated with a 1.7 IQ point decrease in WISC-III performance scores, 1.3 IQ point decrease for WISC-III total scores at age 8.

Fully adjusted models (Model 3) with confounders (maternal age, maternal education, deprivation indices, other two trimesters) and additional covariates (maternal smoking, maternal psychiatric history, gestation, birth weight, and child sex) showed that infections in the third trimester were associated with lower WISC-III performance scores at 8 years (β= - .033, *p*=.032) (Table 3). These results represent a 1.4 IQ point decrease in WISC-III performance IQ scores.

### Sensitivity Analysis

A sensitivity analysis of the study models found consistent associations between infections occurring at the third trimester and outcomes for WISC-III verbal IQ scores (β= - 1.33, *p*=.022), performance IQ scores (β= -1.19, *p*=.028), and total IQ scores (β= -1.39, *p*=.009) at age 8 in the unadjusted model. While no significant associations between occurrence of infections at any trimester and cognitive outcome scores for the partially and fully adjusted models, lower maternal education (*p*<.05) and highest deprivation (*p*<.05) were generally seen to have associations with lowered cognitive scores across all trimesters.

## Discussion

A prospective birth cohort was used to examine associations between maternal infections occurring at each trimester of pregnancy and children’s verbal, performance and total IQ scores at 18 months, 4 years, and age 8. Results from unadjusted models provided evidence for maternal infections occurring during the third trimester being associated with verbal IQ scores at age 4, and verbal, performance, and total IQ scores at age 8. The associations between third-trimester infections and cognitive outcomes were seen to be attenuated after adjustment for confounders, with significant associations seen only for performance and total IQ scores at age 8 after the first adjustment. Further adjustment with additional covariates left only significant associations for performance IQ scores at age 8. A similar prevalence of maternal infections during pregnancy was shown in this study as compared to other cohort samples (Curcio et al., 2020; Harvey & Boksa, 2012).

Children’s IQ outcomes appear to depend on critical periods in gestation, with evidence pointing to effects of infection limited to later gestation. Animal models have provided evidence on potential mechanisms, showing that inflammatory cytokines produced in response to infection may be linked to fetal brain development changes. Mice treated with analogues of bacterial or viral infections at different gestational periods showed different neurodevelopmental profiles of hippocampal reelin and GAD67 cell number expression in the hippocampal dorsal or ventral stratum oriens of offspring (Harvey & Boksa, 2012). Studies investigating infection-associated immunological events on the fetus showed early maternal inflammation to be linked with negative effects on the development of a fetus’ dopaminergic system at multiple levels such as cell distribution and connectivity (Meyer et al., 2007). In contrast, later brain insults during pregnancy have been linked to deficits in cell organization and maturation of synapses which occur over an extended period, affecting cognitive function (Meyer et al., 2006).

While animal models provide a possible explanation as to how brain neurochemistry and structure can potentially be affected by different times of infection exposure, evidence from human studies, especially looking at specific trimester effects and cognitive outcomes, is lacking. Further, while some genetic patterns have been found in maternal intellectual ability and child IQ scores (Lean et al., 2018), no firm conclusions can be drawn that suggests genetics fully accounts for children’s cognitive abilities. Instead, there is current growing emphasis on using the gene-environment interaction to examine the role of maternal inflammation and biological pathways that lead to fetal brain development and cognition (Dozmorov et al., 2018; Rasmussen et al., 2021).

Literature on human studies has been mixed, showing varying results on vulnerability to inflammation occurring in different gestational periods, especially on a child’s cognitive development (Cordeiro et al., 2015; Rosenberg, 2018; Yarnykh et al., 2018). The significant associations between third trimester infection and cognitive outcomes at age 8 identified in this study suggest a possible latent effect of maternal systemic inflammation which may lead to future cognitive deficits. This could possibly be due to an interaction with unaccounted environmental factors after birth, such as lowered socioeconomic status, which could confound child development as it is also considered an inflammatory process during childhood (Schmeer & Yoon, 2016). The link with maternal infections during later gestation possibly shows that despite having fetal brain infrastructure in place, environmental factors such as deprivation could still affect brain structure or functional connections after birth.

This suggests that infections in late pregnancy may influence the postnatal processes of cognitive development such as learning. Cognitive outcomes being affected at a later age could also reflect a possibility of maternal infection effects only becoming evident under increased cognitive load, as later cognitive IQ assessments require more advanced skills such as abstract reasoning.

While this study could not directly infer causality, study results are consistent with previous research on maternal-fetal associations, where clinical evidence show how presence of maternal inflammation during pregnancy using elevated cytokine profiles contribute to prenatal risk programming (Andersson et al., 2016; Wright et al., 2010). Other studies using these same maternal cytokines found negative influences on prenatal central nervous system development (Zaretsky et al., 2004), with animal models showing further evidence for negative neurobehavioral outcomes (Bronson & Bale, 2014). In sum, research supports how prenatal programming consists of multiple variables such as occurrence and regulation of inflammation, genetic susceptibility, and environmental stressors; all of which interact in a complex manner that is still not yet fully understood in humans (Hantsoo et al., 2019), which could explain why study results were mixed. Nevertheless, this study contributes further evidence through the specific scope of how maternal infections during different trimesters of pregnancy could possibly affect childhood IQ outcomes.

### Strengths and Limitations

Main study strengths were the use of a large high quality longitudinal design and a well-characterized sample. While cohort studies such as the ALSPAC tend to have considerable attrition due to reasons such as loss to follow-up related to socioeconomic status, it has been shown that differences in estimates when comparing full or restricted cohorts tended to be small (Howe et al., 2013). Similar infection rates during pregnancy and study attrition allowed for comparability with other cohort studies also looking at mother-child associations. Study limitations include a reliance on mother self-reported infection data; that records of infections were not classified by severity; and that causes of infections (e.g. bacterial/viral/fungal) were not recorded and could not be stratified by pathogen type for analysis. In addition, while the study has included a list of confounders and covariates, not all factors such as parental attitudes or parenting styles and other variables related to the quality of care of the child, or genetic factors could be accounted for in this study. Despite not being able to account for these factors, study results still showed predictive associations, which was consistent with research from both animal and human studies focusing on maternal-fetal interactions. Finally, IQ assessments from the dataset were only measured up to age 8, so lasting effects of cognitive development could not be analyzed beyond those years.

### Implications and Further Study

Our study highlighted possible third-trimester specific effects of maternal infection on children’s cognitive abilities. However, effect sizes were small, suggesting a weak link between maternal infections during the third trimester of pregnancy and childhood cognitive outcomes. One potential direction of future research could include examining mechanisms which mediate these effects of prenatal infections on childhood cognition such as fetal brain gene expression after maternal immune activation (Lombardo et al., 2018). Other research could also explore temporal dependency and specificity to performance IQ scores, whether the effect of infection is moderated by specific early childhood environmental factors, and whether its effect persists in future cognitive outcomes.

## Data Availability

All data produced in the present study are available upon reasonable request to the authors.

## Acknowledgements

We are extremely grateful to all the families who took part in this study, the midwives for their help in recruiting them, and the whole ALSPAC team, which includes interviewers, computer and laboratory technicians, clerical workers, research scientists, volunteers, managers, receptionists and nurse. We further thank the European Union’s Horizon 2020 research and innovation program for their generous support.

